# Norovirus GII wastewater monitoring for epidemiological surveillance

**DOI:** 10.1101/2023.04.10.23288357

**Authors:** ML Ammerman, S Mullapudi, J Gilbert, K Figueroa, FPN Cruz, KM Bakker, MC Eisenberg, B Foxman, KR Wigginton

## Abstract

Norovirus surveillance using case reports and syndromic detection often lags rather than leads outbreaks. To assess the timeliness of norovirus wastewater testing compared with syndromic, outbreak and search term trend data for norovirus, we quantified norovirus GII in composite influent samples from 5 wastewater treatment plants (WWTPs) using reverse transcription-digital droplet PCR and correlated wastewater levels to syndromic, outbreak, and search term trend data. Wastewater HuNoV RNA levels were comparable across all WWTPs after fecal content normalization. Norovirus wastewater values typically coincided with or led syndromic, outbreak, and search term trend data. The best correlations were observed when the wastewater sewershed population had high overlap with the population included by other monitoring methods. The provision of norovirus-specific measures and earlier detection of norovirus found using wastewater surveillance suggests that wastewater-based surveillance of human norovirus GII will enhance existing public health surveillance efforts of norovirus.

## Background

SARS-CoV2 demonstrated the utility of wastewater monitoring of pathogens, and catalyzed the infrastructure development needed to monitor other pathogens of public health importance. One such pathogen is norovirus, which causes more than 90% of epidemic non-bacterial gastroenteritis outbreaks (1). Norovirus is estimated to result in 900 deaths, 109,000 hospitalizations, 465,000 emergency department visits, and 2.3 million clinic visits (2), annually in the United States, causing substantial health and economic burdens (3).

Norovirus, a genera in the *Calciviridae* family, is a nonenveloped virus with a positive-sense RNA genome. There are ten genogroups of Norovirus, GI – GX, and 48 genotypes (4). The GII genogroup, specifically variants of the GII.4 genotype, are the most common cause of norovirus disease worldwide. The most common route of transmission is person-to-person, followed by contaminated food or water, and contact with contaminated fomites (5). An estimated 30% of all norovirus infections and over 40% of GII.4 infections are asymptomatic (6). Reinfection is common due to relatively short-term immunity and the diverse genogroups (7,8). Norovirus is a “winter” pathogen, in the northern hemisphere most outbreaks occur between November and April (9).

Norovirus surveillance practices vary greatly across the US, and there is no requirement for local, territorial, or state agencies to report individual norovirus cases to the national system. Public health officials often rely on syndromic data as early indicators of norovirus outbreaks. Health departments are encouraged to report all waterborne, foodborne, and enteric disease outbreaks, which would include norovirus outbreaks, to the National Reporting System (NORS) and norovirus outbreaks to Calicinet (10). The CDC and health departments from fourteen states, including Michigan, participate in the Norovirus Sentinel Testing and Tracking (NoroSTAT) network. However, the lag between norovirus testing and reporting can be as long as a few weeks - unacceptably long for a highly transmissible virus. Furthermore, data aggregated across participating states has minimal value for local health decisions and much of norovirus transmission goes undetected by conventional surveillance systems due to asymptomatic cases and cases that do not require clinical care.

Wastewater monitoring for norovirus has the potential to provide more local, early-warning information to inform public health decision-making—potentially prior to clinically detected outbreaks. Human norovirus is detectable in wastewater treatment plant (WWTP) influent (11–13) and studies have reported positive correlations between wastewater levels and conventional surveillance data, including confirmed HuNoV cases (14–16), GI illness cases (17,18), and confirmed hospital cases (18,19). A study in Japan found stronger correlations between wastewater levels and regional rather than national cases (17). Wastewater data led conventional surveillance data in some reports and not in others. These discrepancies could be due to a number of reasons, including that the regions covered by the sewershed did not always correspond to the regions covered by the conventional surveillance data, differences between the study populations, the different types of surveillance data used between the studies (e.g., hospital HuNoV cases, confirmed GI cases, syndromic data, etc.), and the low temporal resolution of wastewater and conventional surveillance data used in some of the studies. This suggests a need to examine corresponding wastewater and epidemiological surveillance data for multiple populations at high resolution.

Here, we measure HuNoV GII levels at high resolution in five WWTPs over one year. We focus on HuNoV GII testing because 392 of the 440 US confirmed norovirus outbreaks recorded by Calcinet (or 89.1%) in 2021-2022 were HuNoV GII. We demonstrate that, like SARS-CoV-2, wastewater surveillance of HuNoV would be valuable for public health surveillance, particularly in combination with other traditional epidemiological surveillance methods, and has the benefit of not relying on clinical testing or school, hospital, or health department reporting.

## Methods

The Michigan SARS-CoV-2 Epidemiology - Wastewater Evaluation and Reporting (SEWER) Network includes local partnerships between wastewater utilities, health departments, tribal communities, universities, and laboratories that are collecting and analyzing wastewater for SARS-CoV-2 across the state of Michigan. All SEWER network laboratories use influent samples and approximately the same RNA purification methods. This infrastructure allowed us to monitor samples for additional RNA viruses.

### Sample collection procedure

WWTP personnel collected daily influent samples from five WWTPs in southeast Michigan (Ann Arbor, Flint, Jackson, Tecumseh, and Ypsilanti; Table 1) between 7/18/2021 and 7/14/2022, except Tecumseh where sample collection began 1/12/2022. Samples were collected by 24-hour composite samplers kept at 4 °C, and 50 ml aliquots were delivered biweekly by courier on ice to the University of Michigan. Samples were stored at 4°C and processed within 120 hours of collection.

**Table 1.** Information about wastewater catchment areas. The sewershed population data for Ann Arbor and Flint WWTPs was obtained from the 2022 American Community Survey 5 year data, and the State of Michigan SWEEP website was the source for Jackson, Tecumseh, and Ypsilanti Community WWTPs. https://www.michigan.gov/coronavirus/stats/wastewater-surveillance/dashboard/sentinel-wastewater-epidemiology-evaluation-project-sweepwebsite.

| Wastewater Treatment Plant (WWTP) | Abbreviation | Counties Covered by WWTP | Sewershed Population | Combined/ separate |
| --- | --- | --- | --- | --- |
| Ann Arbor | AA | Washtenaw | 121,093 | separate |
| Flint | FL | Genesee | 95,999 | combined |
| Jackson | JS | Jackson | 90,000 | separate |
| Tecumseh | TM | Lenawee | 8,680 | separate |
| Ypsilanti Community | YC | Washtenaw and Wayne | 330,000 | separate |

### Sample processing

We concentrated viruses in 40 ml samples 8-40 fold using PEG precipitation as previously described (20). RNA extraction was performed with 200ul of sample concentrate using the QIAmp Viral RNA Mini Kit (Qiagen Sciences, MD) with an elution volume of 80ul of water. A single extraction was performed for each sample collected. BCoV (Bovilis Coronavirus Vaccine, Merck Animal Health, NJ) was added to samples prior to RNA extraction reactions as a recovery control. Negative extraction controls and positive BCoV and Norovirus GII extraction controls were also prepared. Pepper Mild Mottle Virus (PMMoV) was quantified as a measure of fecal content (21).

### RT-ddPCR

Reverse transcription - digital droplet PCR (RT-ddPCR) analysis of Norovirus GII was performed on RNA samples using multiplexing (with SARS N1 and N2) or triplexing (with BCoV and PMMoV). Norovirus concentrations were normalized by PMMoV concentrations to account for fecal content. Assay details as per MIQE guidelines (22) are provided in the Appendix.

### Syndromic, Outbreak, and Internet Search Term Data

School reported GI illnesses and emergency department GI related visit data were provided by local public health departments. These syndromic data included only reports of symptoms associated with norovirus and no tested or confirmed norovirus cases. Per the CDC National Center for Health Statistics standard, to prevent disclosure of individual information, only data with > 10 cases was included. Weekly total norovirus outbreak values from September 25, 2021 through June 12, 2022 were obtained from the CDC NoroSTAT website (www.cdc.gov/norovirus/reporting/norostat) which compiles and reports outbreak data from 14 states including Michigan, representing about 25% of the US population.

Google Trends records how often a term is searched for in a given region relative to its total search volume with the normalized value in a range from 0 to 100. We obtained time-series data for the search terms “norovirus”, “gastroenteritis”, “stomach flu” for the Metropolitan Detroit region and the state of Michigan from 1/1/2021-11/29/2022. Data were joined by wastewater sample collection date and search term data week. Search term values were rolled forward to apply to all dates in a given week for matching purposes.

### Statistical Analyses

We conducted statistical analyses with Graphpad Prism 9.4.1 except where otherwise noted. We determined the mean, median, and interquartile range of log-transformed weekly average HuNoV GII wastewater concentrations (gc/L) and the log-transformed weekly average HuNoV GII/PMMoV wastewater concentration ratios from each WWTP, from all WWTPs combined, and by season.

We calculated Pearson correlations and cross correlations between wastewater HuNoV GII values and the number of school-reported GI illnesses, emergency department GI visits, and weekly total outbreaks from NoroSTAT. Negative cross correlation lag times indicate clinical data leads wastewater data and positive values indicate wastewater values lead (See Appendix for details).

Pearson correlations and cross correlations were also computed between HuNoV GII against Detroit and Michigan-wide search term trends using R version 4.0.3 (2020-10-10) -- “Bunny-Wunnies Freak Out”; Copyright (C) 2020 The R Foundation for Statistical Computing; Platform: x86_64-w64-mingw32/x64 (64-bit).

## Results

### HuNoV GII RNA levels and trends for the five individual WWTPs and all WWTPs combined

Wastewater HuNoV RNA levels were highest in the winter and spring, and lowest in the summer and fall (Figure 1). Including all 5 WWTPs, the daily values ranged from 6 × 10^3^ gc/L to 9.7 × 10^7^ gc/L and the median weekly value was 6.0 log_10_ gc/L (Supplementary Figure 1). Although TM had the highest overall values, TM data was only collected between January-July 2022. If Tecumseh was excluded the median value is 5.96, with an interquartile range (IQR) of 5.46-6.43. FL, the only sewershed with a combined (stormwater and sanitary) system, also had a median HuNoV RNA value that fell outside of the IQR of all WWTP data.

**Figure 1.**
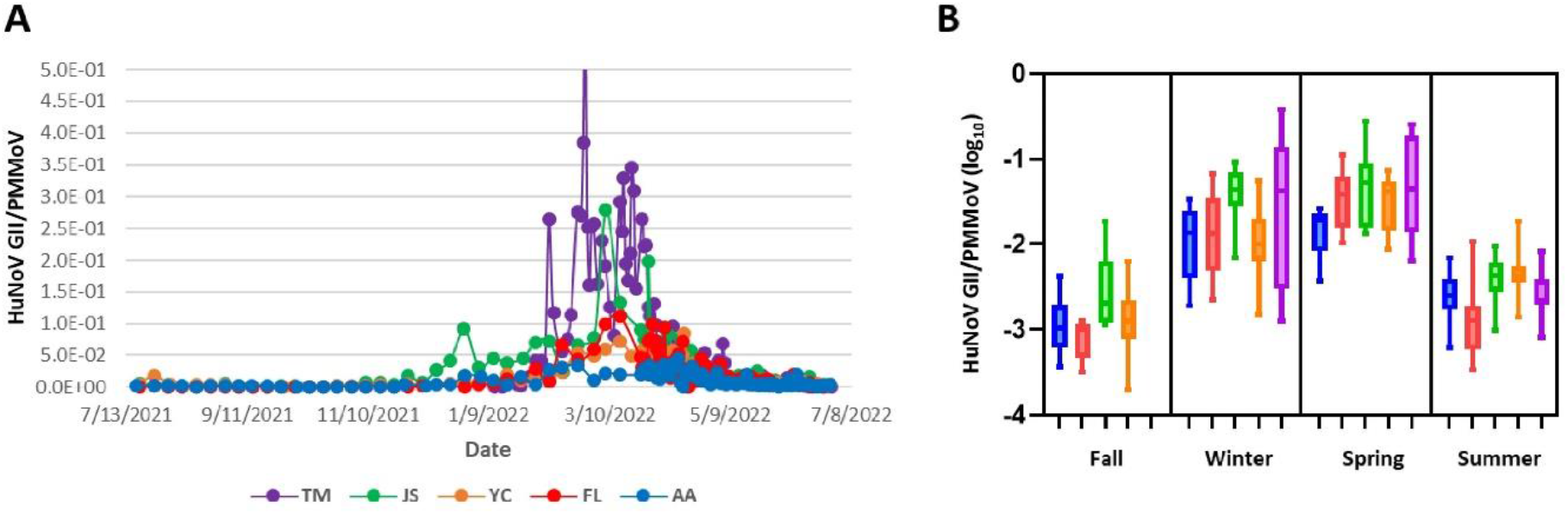
HuNoV GII wastewater levels at 5 wastewater treatment plants in 2021-2022. A. Comparison of PMMoV-normalized HuNoV GII levels from 5 WWTPs in Michigan from 2021-2022. HuNoV was quantified in influent samples using ddPCR at least weekly and PMMoV normalized values were plotted over one year. The only exception is TM, where sample collection began later, in January 2022. Lines connect all values measured for each WWTP. B. Seasonal HuNoV GII values in log10 (median, IQR) for each individual WWTP are presented using box-and-whisker plots for all 4 seasons. Note, no fall values for TM were obtained. The definition of the seasons is meteorological, beginning on the 1^st^ day of the months of the equinoxes or solstices.

To account for variations in fecal content, we normalized HuNoV GII values to PMMoV values. The general patterns over the year were similar using either HuNoV gc/L concentrations or PMMoV-normalized concentrations (Supplementary Figure 1 and Figure 1A). The PMMoV-normalized average weekly values correlated strongly with the average weekly HuNoV gc/L values for samples from each WWTP, with correlation coefficients between 0.72 (AA WWTP) to 0.83 (TM WWTP) (Supplementary Figure 2). We focus on PMMoV-normalized HuNoV data for the remainder of the manuscript but include the HuNoV GII values in gc/L in the Supplementary Material to enable direct comparison with previously published data.

When data from the five WWTPs were combined, the highest HuNoV RNA values were obtained in the spring (average values of 0.027 HuNoV GII/PMMoV or 3.89 × 10^6^ gc/L) and the lowest in the fall (0.0012 HuNoV GII/PMMoV or 2.57 × 10^5^ gc/L), (Figure 1B, Supplementary Figures 3A and 4A). In general, fall and summer HuNoV RNA levels were similar and the winter and spring HuNoV RNA levels were similar for all five WWTPs (Figure 1B, Supplementary Figures 3 and 4). The HuNoV RNA concentration variations observed within a plant also exhibited seasonal trends. Specifically, the Interquartile ranges (IQRs) observed in winter and spring for a specific plant were larger than the IQRs observed in summer and fall for most WWTPs (Figure 1B).

### Comparison of HuNoV GII wastewater levels to School GI Illnesses and GI-related Emergency room visits

#### School GI Illnesses

We compared HuNoV GII levels measured in TM, AA, and YC wastewater to reported weekly school gastrointestinal illness numbers (Figure 2). Wastewater values and school GI illnesses correlated for TM (Figure 2A), AA and YC WWTPs (Figure 2B). For TM, the obtained school GI numbers are for the town of Tecumseh, whereas for AA and YC, the obtained GI numbers are for Washtenaw County. Of note, the AA WWTP sewershed is entirely included within Washtenaw County, whereas only one-third of the YC WWTP sewershed population is within Washtenaw County (the remainder in Wayne County). Due to the large variations in Washtenaw County school attendance, we normalized GI reports for Washtenaw County per 1000 students in attendance (details in Appendix).

**Figure 2.**
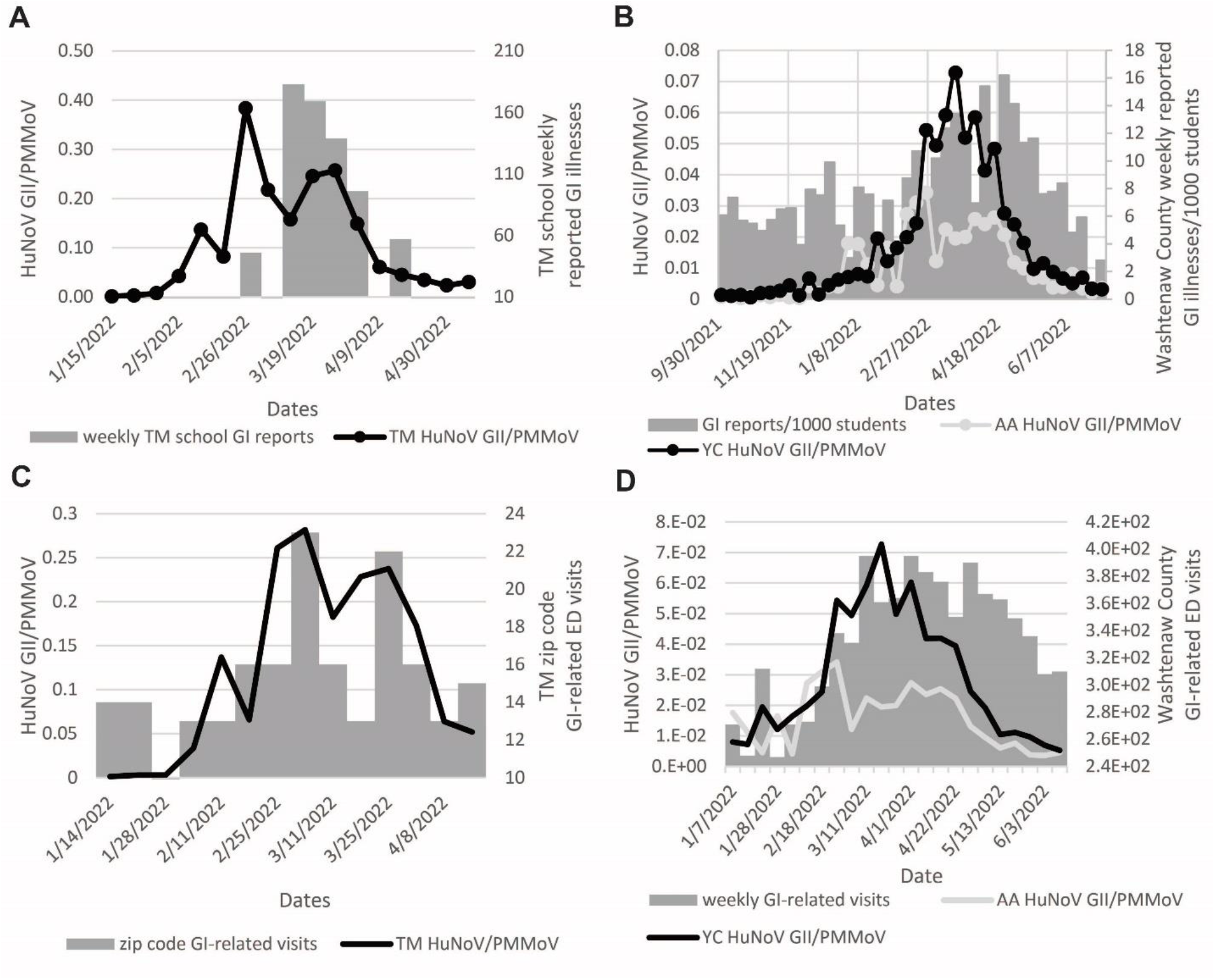
Correlation of TM, AA, and YC HuNoV GII wastewater values with syndromic data. A. TM WWTP HuNoV GII/PMMoV values along with weekly school reported gastrointestinal illnesses at TM schools. B. AA and YC wastewater levels as HuNoV GII/PMMoV and county-level school reported GI illnesses normalized to enrollment. The GI reports per 1000 people were calculated to normalize for large variations in school attendance. C. Correlation between the weekly total number of zip code level GI-related ED visits and TM PMMoV-normalized HuNoV GII wastewater values. D. Correlations between the weekly total number of county-level GI-related ED visits and AA and YC PMMoV-normalized HuNoV GII wastewater values.

#### Wastewater HuNoV RNA concentrations led reported GI cases

To determine the relative timing of wastewater HuNoV values and school GI illnesses, we used a temporal cross-correlation for weekly total school reported GI illnesses from -2 to +3 or +5 weeks relative to the weekly average HuNoV GII/PMMoV wastewater values. Cross-correlation of TM wastewater and GI case data showed a significant correlation on the same day (lag = 0, r = 0.6), 1 week (+1) later (r = 0.67), and 2 weeks (+2) later (r = 0.82), (Table 2). For AA, very high correlations were observed for +1 to +4 weeks, with p values <0.0001 (Table 2), with the highest correlation for +3 and +4 weeks (r= 0.67 and 0.66). For YC, correlation coefficients greater than 0.65 (p values <0.0001) were observed for all dates from +1-4 weeks later (Table 2) and the highest correlation was observed at +3 weeks (r = 0.74). The same general correlation trends were observed for each WWTP when HuNoV GII concentrations (gc/L) were used in the correlation analysis instead of PMMoV-normalized values (Supplementary Figure 5).

**Table 2.**
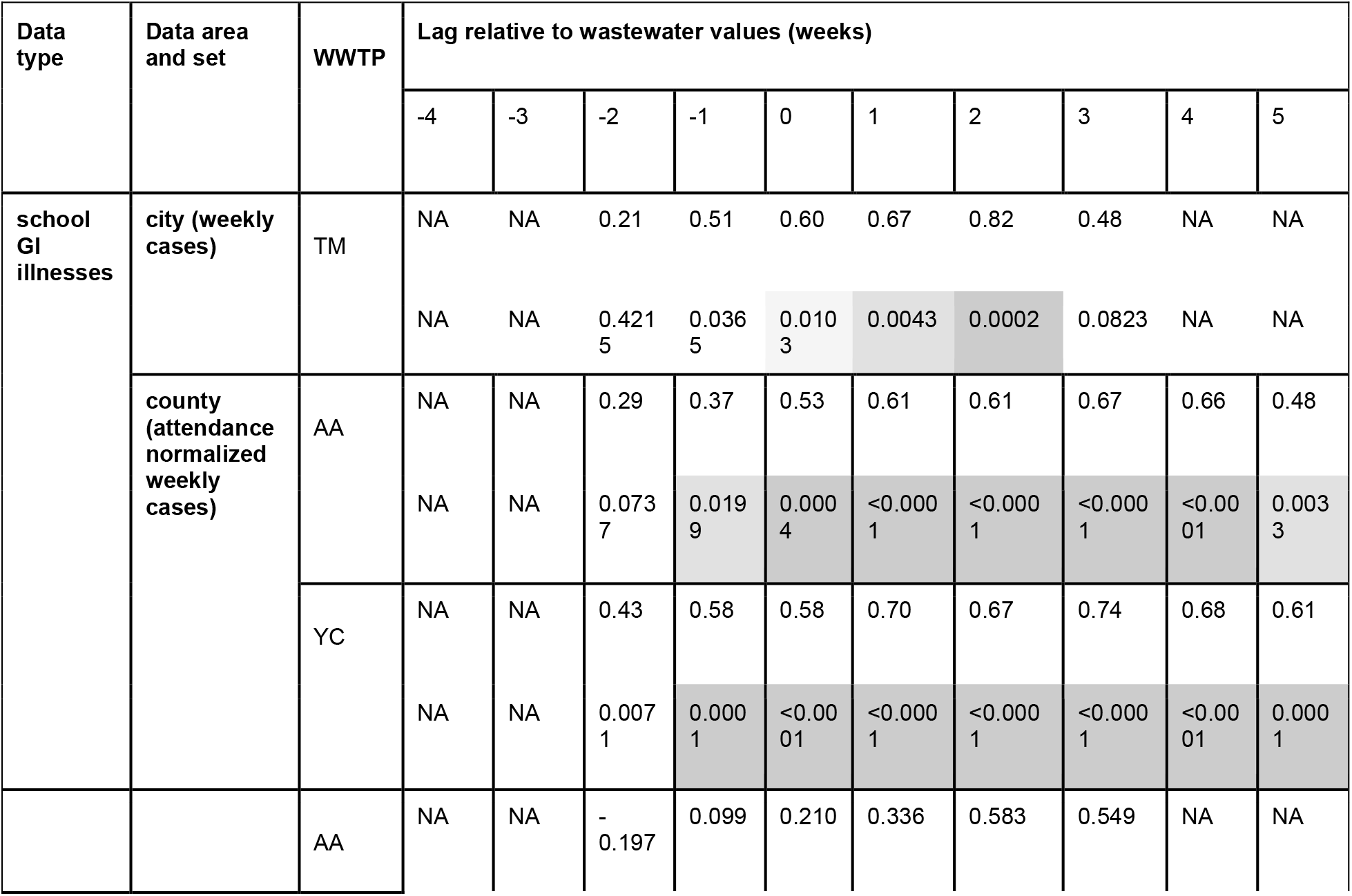
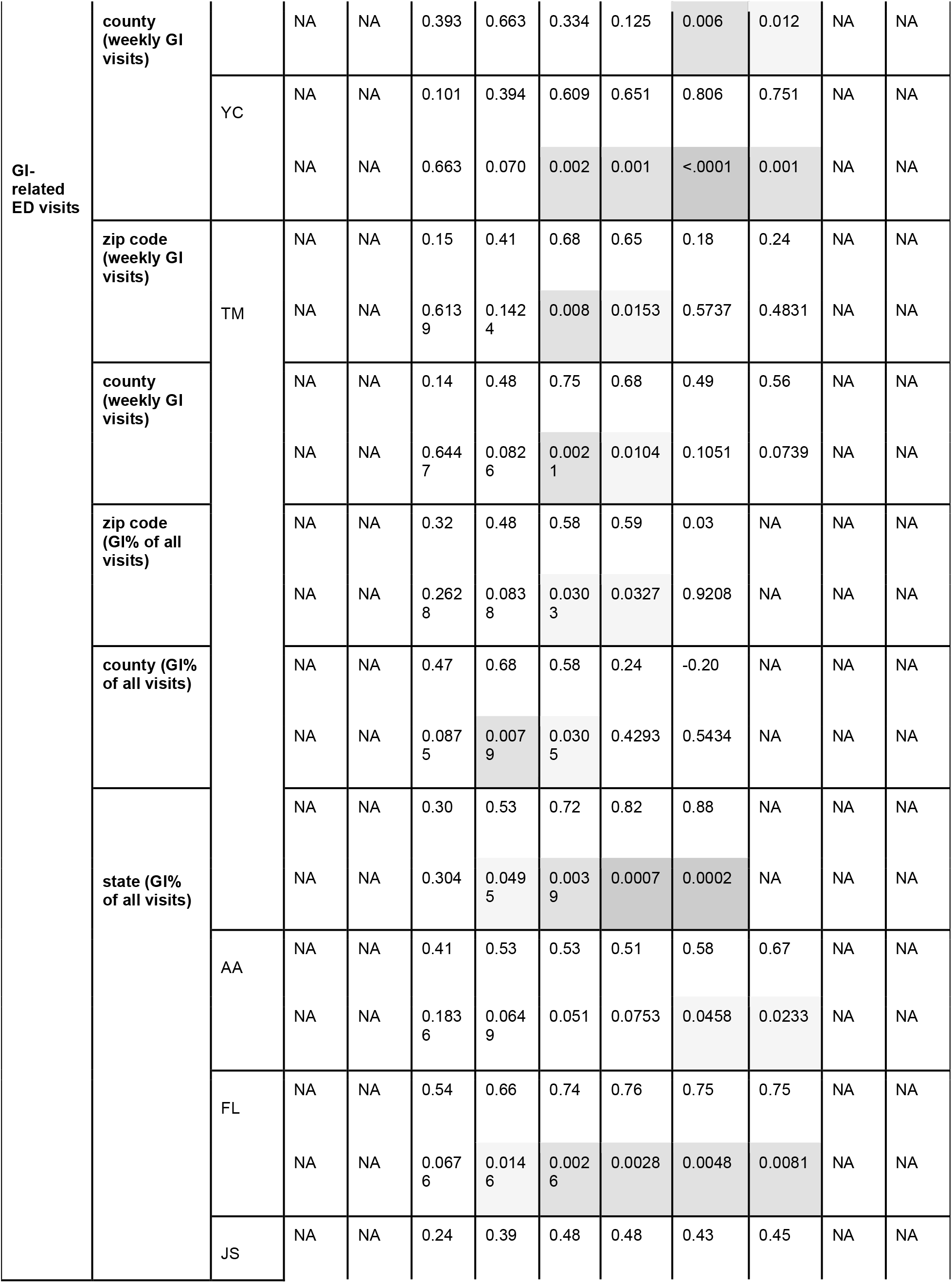
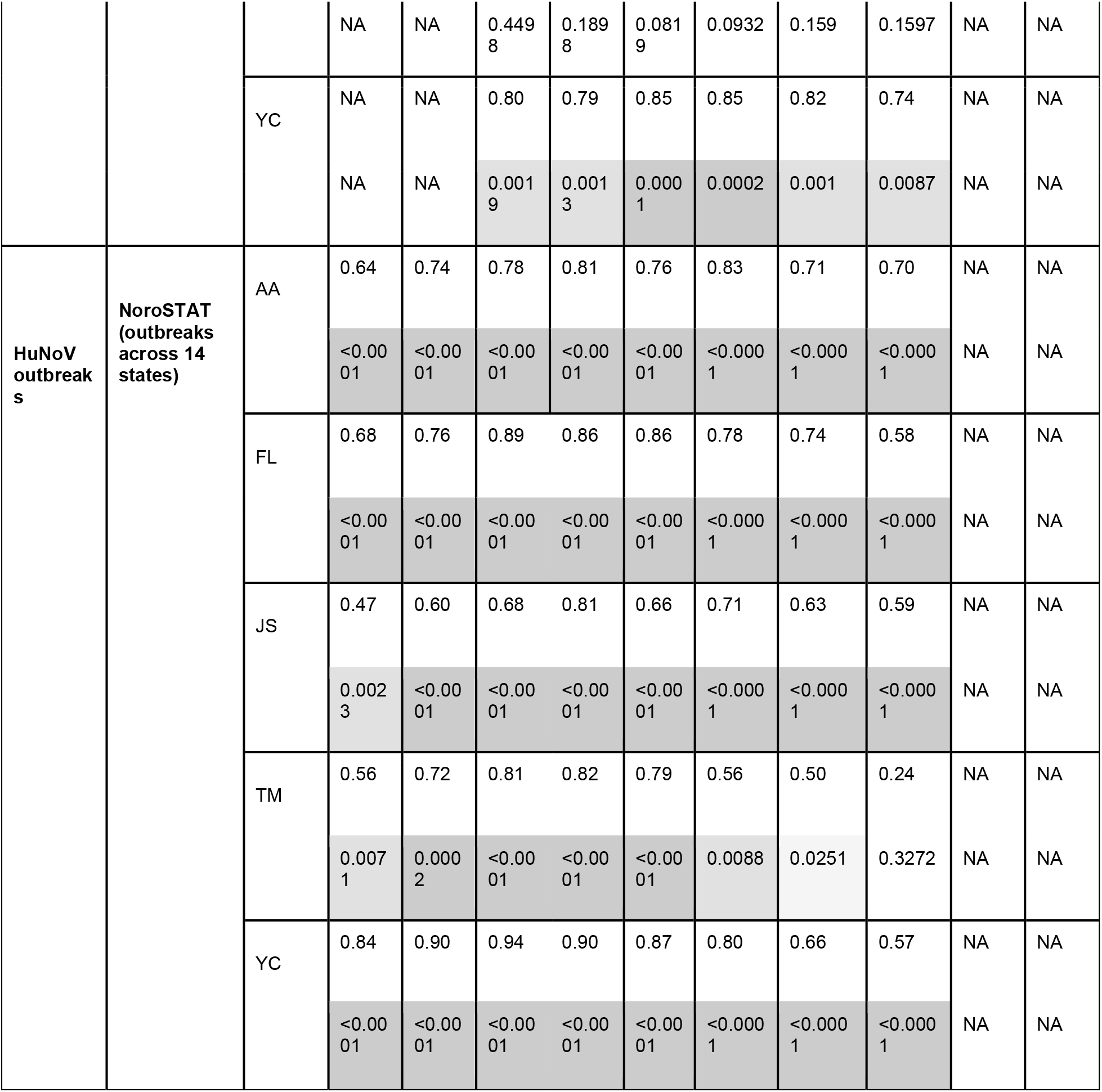
Cross correlations of HuNoV GII wastewater values with conventional monitoring data. For each data set the top row is the Pearson correlation coefficient (r) and the bottom row is the p value. The p values have been color coded with light gray for p < 0.05, medium gray for p < 0.01, and dark gray for p < 0.001.

#### GI-related emergency department visits

HuNoV GII wastewater levels from TM were compared to the total number of GI-related ED visits in the respective zip code (Figure 2C and Table 2) and county (Table 2), and to the percent of ED visits that were GI-related in the respective zip code and county. We observed a significant correlation between GI-related ED visits and average weekly HuNoV wastewater values for the same day (r= 0.68) and +1 week (r=0.65, wastewater leading), with the highest correlation values for the same day when comparing the total number of GI-related ED patients from the TM zip code to the weekly average TM HuNoV GII/PMMoV wastewater values (Table 2). The same trend was seen when comparing TM HuNoV GII/PMMoV wastewater values to the total number of ED patients from Lenawee County with GI symptoms, with R values of 0.75 and 0.68 for 0 to +1 week (Table 2).

Cross correlation analyses between the percent of ED visits in the TM zip codes that were GI-related and the TM HuNoV GII/PMMoV wastewater values found the highest correlation occurred for the same day (R=0.58) and +1 week (R= 0.59), similar to what was seen when comparing total patient number (Table 2). The highest correlation for county percent ED data was at -1 week (R= 0.68) with a statistically significant value also seen on the same day (R= 0.58). This is a slight shift compared to results using total patient data. The TM wastewater values had significant correlation with the percentage of GI-related ED visits across the state of Michigan from -1 to +2 weeks, with the highest correlation seen at +2 weeks (R=0.88) (Table 2).

For AA and YC, HuNoV GII wastewater levels were compared to total weekly GI-related ED visits in the county. Similar to TM, we observed a significant correlation between total weekly GI-related ED visits in Washtenaw County and average weekly AA and YC HuNoV GII/PMMoV values (Figure 2D). The highest correlation values were obtained at +2 weeks for both WWTPs (AA R= 0.58; YC R= 0.81), and significant correlations were observed with AA for +3 weeks and with YC for 0, +1, and +3 weeks (Table 2). We also correlated the HuNoV GII wastewater values from AA, YC, Jackson (JS), and Flint (FL) WWTPs to Michigan state ED GI-related visit values. We saw significant correlations for all WWTPs, except JS. The timing of the highest correlations varied across the five WWTPs, although wastewater was consistently the leading indicator, with AA +3 weeks (R= 0.67), FL +1 week (R= 0.76), TM +2 weeks (R= 0.88), YC +1 week (R= 0.85), and JS +1 week (R= 0.48) (Table 2). Most WWTPs had extended time periods over which a significant correlation was seen, e.g., YC HuNoV wastewater values had significant correlation with state syndromic data from -2 to +3 weeks (Table 2).

Overall, similar cross-correlation results were observed for ED data when wastewater HuNoV GII values in gc/L were used instead of HuNoV GII/PMMoV (Supplementary Figures 5 and 6). In some cases, correlations were poorer with gc/L; e.g., AA data resulted in no significant correlations.

### NoroSTAT outbreak numbers

The temporal patterns for the 2021-2022 norovirus outbreaks reported by NoroSTAT and HuNoV GII wastewater values were similar (Figure 3A). Wastewater data from the 5 WWTPs correlated well with NoroSTAT data (R= 0.50 to 0.94) for each WWTP from -3 to +2 weeks when comparing the timing of the outbreaks to the HuNoV GII wastewater levels (Figure 3B and Table 2). The highest correlation occurred from -2 to +1 weeks, depending on the WWTP.

**Figure 3.**
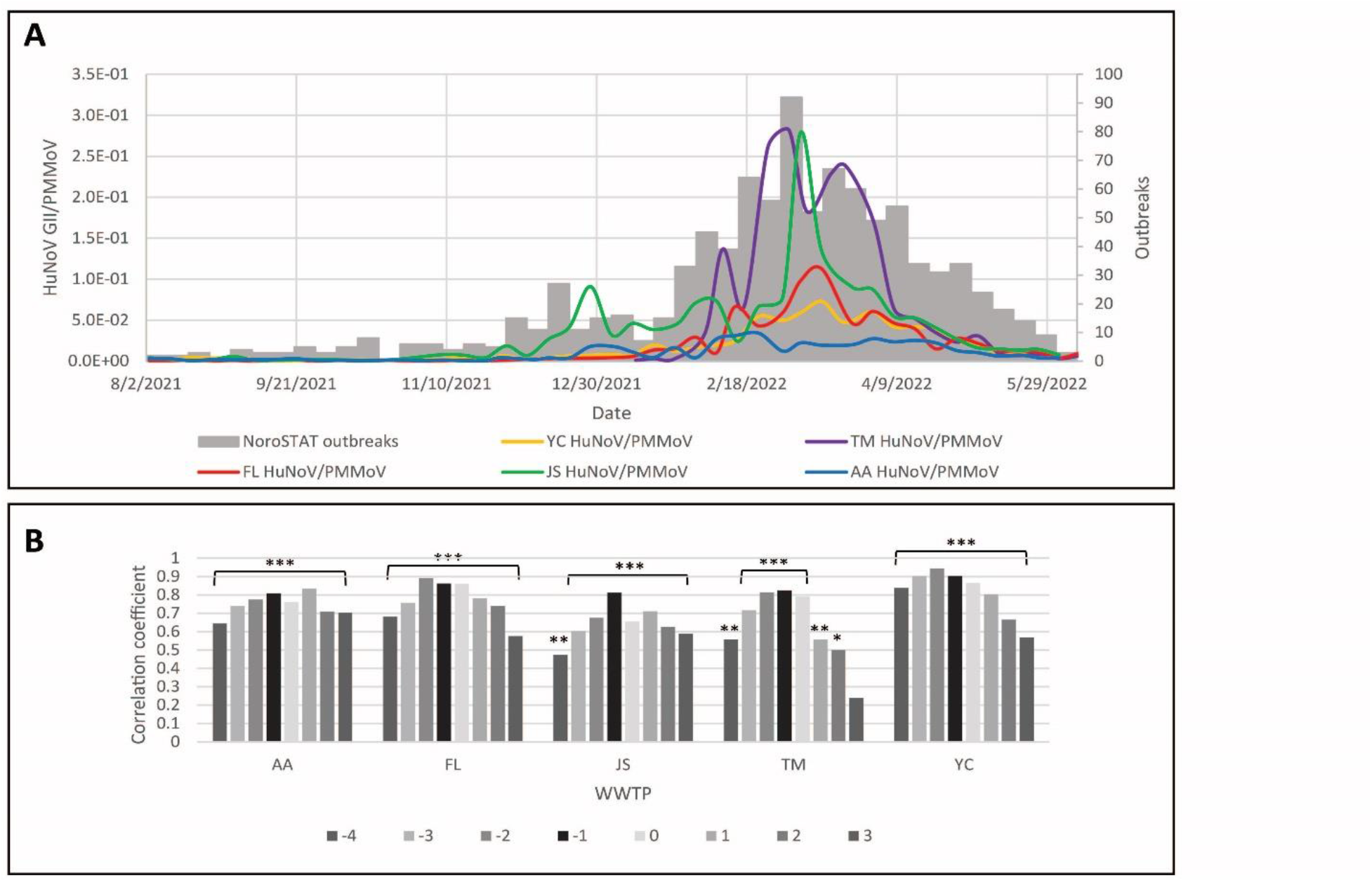
Correlation of HuNoV GII wastewater values with NoroSTAT reported outbreaks. A. Time course from August 2021 to June 2022 showing weekly average HuNoV GII/PMMoV values from 4 WWTPs and weekly norovirus outbreaks reported by NoroSTAT. TM WWTP data is only from January –June 2022. B. Cross correlations between NoroSTAT outbreak numbers and weekly average HuNoV GII/PMMoV values. Graph showing Pearson’s correlation coefficients (r) and probability values (p) determined by comparing weekly average HuNoV GII/PMMoV wastewater values from all 5 WWTPs to the weekly total outbreaks reported by NoroSTAT. Cross correlations were tested for a lead (−4 to -1 weeks), same timing (0), or a lag (1 to 3 weeks) in outbreak reports compared to wastewater values. Note: * = p<0.05, ** = p<0.01, *** = p<0.001

### Search term data

We compared the number of weekly internet searches for terms related to GI illnesses in Michigan and the Detroit metropolitan (metro) area to HuNoV GII wastewater levels at our five WWTPs. An increase in “Norovirus” and “stomach flu” searches in early 2022 appeared to coincide with increasing wastewater norovirus levels (Figure 4A shows YC as an example).

**Figure 4.**
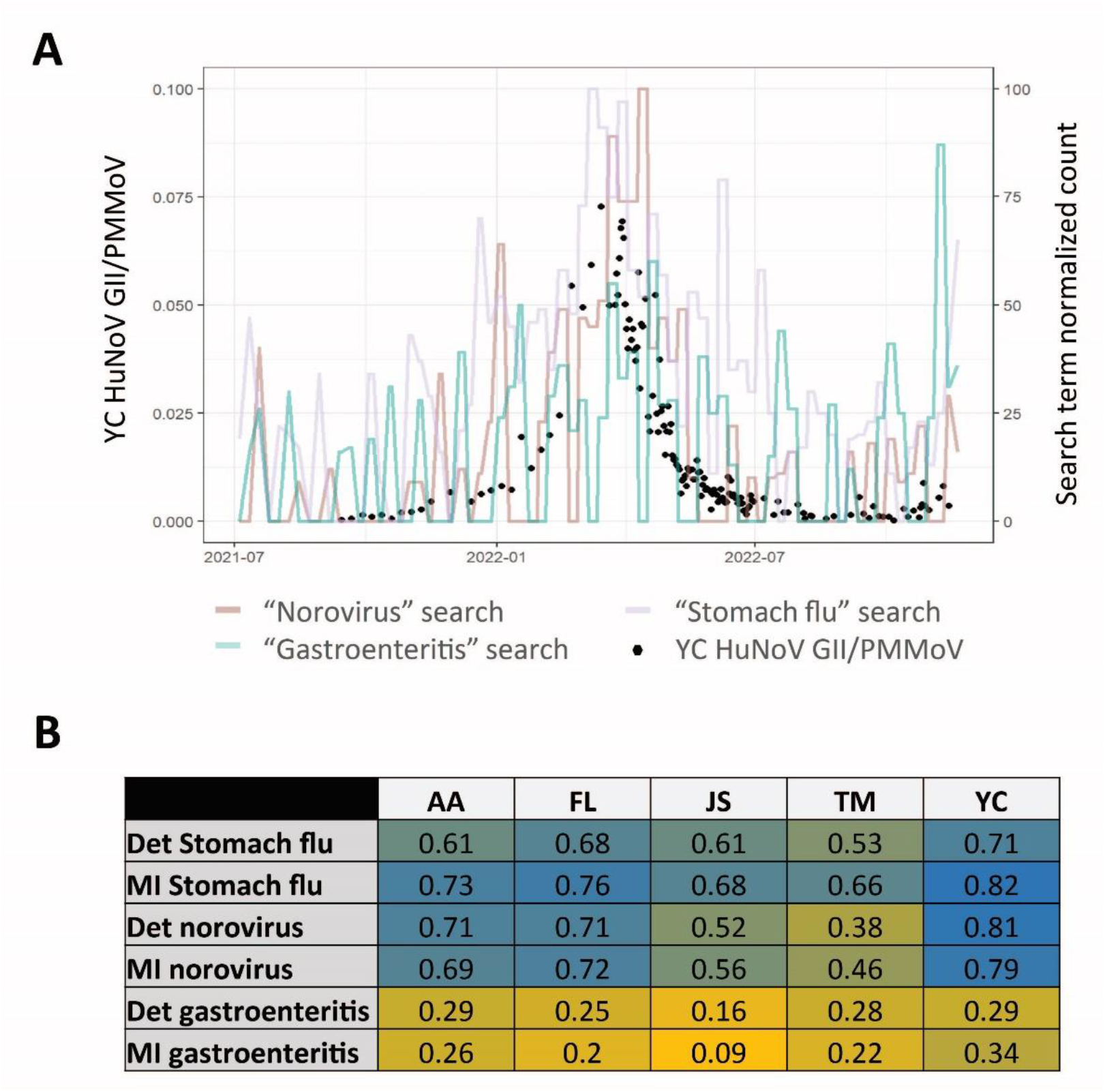
Correlations of HuNoV GII wastewater values with Norovirus-related search terms. A. Time course showing daily YC HuNoV GII/PMMoV values (black dots) and normalized search term trends for the norovirus-related terms “norovirus” (orange), “stomach flu” (lavender), and “gastroenteritis” (green) for the Detroit Metro region. B. Cross correlations comparing HuNoV GII/PMMoV values from all 5 WWTPs and search term trends for “stomach flu”, “norovirus”, and “gastroenteritis” in the state of Michigan and the Detroit (Det) metro area. Pearson’s correlation coefficients (r) are displayed in a heat map with higher values in blue and lower values in yellow.

Further, “stomach flu” in the Detroit metro area and Michigan had positive associations with HuNoV GII wastewater values at all 5 WWTPs, ranging from 0.53 to 0.71 for the Detroit metro area and 0.66 to 0.82 for the state of Michigan (Figure 4B). Four of the 5 plants showed similarly high correlations for the search term “norovirus”. The smallest WWTP, TM, showed lower values (0.38 for the Detroit metro area, 0.46 for MI), potentially due to the small population and earlier timing of local outbreaks (Figure 4B). The highest correlation values were seen for the largest WWTP, YC, which also has its sewershed population in the Detroit metro area included in the google trend search (Supplementary Figure 5). Cross correlations showed variations in timing for HuNoV GII wastewater concentrations compared to search term trends, with the majority, but not all, showing wastewater values leading search term trend data (Supplementary Figures 8 and 9).

We did not see as high correlations for “gastroenteritis” search trends as we did with the search trends for the other terms or with HuNoV GII wastewater values (Figure 4B and Supplementary Figure 10), highlighting the importance of terminology.

## Discussion

In this study we assess the comparability of high spatiotemporal resolution HuNoV GII levels in wastewater from 5 WWTPs, to syndromic, outbreak and search trend data. Norovirus wastewater values typically coincided with or led all other epidemiological data sources, but correlations between wastewater and other data sources varied by the degree of overlap between the sewershed and the population catchment of the other data source. When correlations were made between city and county-level school reported GI illnesses, we observed a 2-3 week delay in the peak school reported GI illnesses compared to the peak of HuNoV GII wastewater levels. WWTPs varied by size of sewershed and population, but after normalization using PMMoV as a fecal indicator, levels were comparable across all WWTPs. Seasonal trends were consistent with previous reports and showed strong seasonal differences across two orders of magnitude, perhaps partially explaining previous observations of high variation in influent norovirus RNA levels across studies and over time (11,13), as previous studies typically used lower temporal resolution and shorter time series. Our study adds to the existing literature by considering: multiple treatment plants spanning a range of population density and urbanicity, plants based in the US, high resolution time series data spanning over a year from 600+ wastewater samples, and comparing wastewater data to multiple epidemiological data sets—including taking a digital epidemiology approach (23).

We detected a large range of HuNoV GII concentrations across all WWTPs, with similar mean and median values across sites. The normalization for fecal content is a strength and a limitation; it enabled comparison across plants but could introduce error. Although we used PMMoV for normalization, other normalization methods including crAssphage, HF183, total nitrogen, or total phosphorus were not assessed and could be as or more effective. We compared wastewater HuNoV GII values to school and hospital syndromic case numbers, NoroSTAT outbreak reports, and norovirus-related search term trends. A limitation of this and future studies is the lack of gold standard case data for norovirus which required triangulation across multiple epidemiological datasets to validate wastewater detection. Additional limitations include that several of our epidemiological data sets could not be resolved at the same spatial geography as our wastewater sewershed data (for example, the Google Trends data could only be resolved to the Detroit region rather than the catchment areas), potentially reducing correlations due to differences in the populations measured, and biasing our lead/lag times if norovirus spatial spread patterns led to one population experiencing increases in transmission patterns before the other population did.

The largest variation in HuNoV GII concentration was detected at our smallest WWTP, which is consistent with previous work reporting wastewater plants with smaller sewershed populations having a higher sensitivity to infection clusters (24). Data sets that aggregate large populations (e.g. the state ED GI visits) aggregate many localized outbreaks that broadly occur seasonally but not at precisely the same times—leading to these data sets tending towards a broader, more hill-like peak. By contrast, data sets representing smaller population sizes may be more likely to discern individual or clusters of outbreaks, which for norovirus can be quite rapid/explosive (25), leading to sharper, more ‘spiky’ increases.

Our results suggest that smaller populations and closer overlap between the wastewater and syndromic or case populations result in closer temporal correlation. However, there is a limit to how small a sewershed population can be before other factors such as large variability in signals and individual variations in shedding become an issue (26), and this may vary by pathogen. Additionally, generally wastewater data drawn from a larger population catchment area tended to have broader peaks (similar to what was observed for syndromic and outbreak data), likely due to the larger number of outbreaks being aggregated across the population in the data.

In summary, our results suggest that wastewater monitoring of norovirus leads or concurs with other epidemiological monitoring methods. The advantages are the more timely availability of wastewater data compared to syndromic and case data, and detection of asymptomatic and mild cases. However, these benefits depend on testing frequency, timely reporting, and diversity and representation of the WWTPs being tested. Given that wastewater data for norovirus correlated closely with multiple other syndromic, outbreak, and search term measures of norovirus activity, and did so either concurrently or as a leading indicator, when appropriately implemented wastewater surveillance of norovirus can provide a useful early warning system and an alternative data stream with which to triangulate norovirus patterns - one that does not require healthcare seeking, clinical testing, or inference based on symptom patterns.

## Data Availability

All data produced in the present work is either contained in the manuscript or available upon reasonable request to the authors.

We thank the Ann Arbor WWTP, Flint WWTP, Jackson WWTP, Tecumseh WWTP, and Ypsilanti Community WWTP, Lenawee County Health Department, Jackson County Health Department, Genesee County Health Department, and Washtenaw County Health Department, especially Kristen Schweighoefer, Laura Bauman, and Juan Marquez.

This study was supported by funding from the University of Michigan through the Public Health Infection Prevention and Response Advisory Committee (PHIPRAC), and from the Michigan Department of Health & Human Services through the Michigan Sequencing Academic Partnership for Public Health Innovation and Response (MI-SAPPHIRE) grant (S MA-2022 (ELCEDE-UM) 10/01/2021 – 07/31/2024, BF mPI) and wastewater surveillance program (SEWER network grant, KW and ME co-PIs).

## References

1. Chachu KA, LoBue AD, Strong DW, Baric RS, Virgin HW. Immune mechanisms responsible for vaccination against and clearance of mucosal and lymphatic norovirus infection. PLoS Pathog. 2008;4(12):e1000236.

2. Burke RM, Mattison CP, Pindyck T, Dahl RM, Rudd J, Bi D, et al. Burden of Norovirus in the United States, as estimated based on administrative data: Updates for Medically Attended Illness and Mortality, 2001–2015. Clinical Infectious Diseases. 2021;73(1):e1–8.

3. Cannon JL, Bonifacio J, Bucardo F, Buesa J, Bruggink L, Chi-Wai Chan M, et al. Global trends in norovirus genotype distribution among children with acute gastroenteritis. Emerg Infect Dis. 2021;27(5).

4. Chhabra P, de Graaf M, Parra GI, Chan MCW, Green K, Martella V, et al. Updated classification of norovirus genogroups and genotypes. J Gen Virol. 2019;100(10):1393.

5. De Graaf M, Van Beek J, Koopmans MPG. Human norovirus transmission and evolution in a changing world. Vol. 14, Nature Reviews Microbiology. 2016.

6. Miura F, Matsuyama R, Nishiura H. Estimating the asymptomatic ratio of norovirus infection during foodborne outbreaks with laboratory testing in japan. J Epidemiol. 2018;28(9).

7. Shah MP, Hall AJ. Norovirus Illnesses in Children and Adolescents. Vol. 32, Infectious Disease Clinics of North America. 2018.

8. Donaldson EF, Lindesmith LC, LoBue AD, Baric RS. Viral shape-shifting: norovirus evasion of the human immune system. Nat Rev Microbiol. 2010;8(3):231–41.

9. Yen C, Wikswo ME, Lopman BA, Vinje J, Parashar UD, Hall AJ. Impact of an Emergent Norovirus Variant in 2009 on Norovirus Outbreak Activity in the United States. Clinical Infectious Diseases. 2011;53(6).

10. Vega E, Barclay L, Gregoricus N, Williams K, Lee D, Vinjé J. Novel surveillance network for norovirus gastroenteritis outbreaks, United States. Emerg Infect Dis. 2011;17(8).

11. Eftim SE, Hong T, Soller J, Boehm A, Warren I, Ichida A, et al. Occurrence of norovirus in raw sewage–a systematic literature review and meta-analysis. Water Res. 2017;111:366–74.

12. Kitajima M, Cruz MC, Williams RBH, Wuertz S, Whittle AJ. Microbial abundance and community composition in biofilms on in-pipe sensors in a drinking water distribution system. Science of the Total Environment. 2021;766.

13. Huang Y, Zhou N, Zhang S, Yi Y, Han Y, Liu M, et al. Norovirus detection in wastewater and its correlation with human gastroenteritis: a systematic review and meta-analysis. Vol. 29, Environmental Science and Pollution Research. 2022.

14. Kazama S, Masago Y, Tohma K, Souma N, Imagawa T, Suzuki A, et al. Temporal dynamics of norovirus determined through monitoring of municipal wastewater by pyrosequencing and virological surveillance of gastroenteritis cases. Water Res. 2016;92.

15. Rajko-Nenow P, Waters A, Keaveney S, Flannery J, Tuite G, Coughlan S, et al. Norovirus genotypes present in oysters and in effluent from a wastewater treatment plant during the seasonal peak of infections in ireland in 2010. Appl Environ Microbiol. 2013;79(8).

16. Wang H, Neyvaldt J, Enache L, Sikora P, Mattsson A, Johansson A, et al. Variations among Viruses in Influent Water and Effluent Water at a Wastewater Plant over One Year as Assessed by Quantitative PCR and Metagenomics. Appl Environ Microbiol. 2020;86(24).

17. Kazama S, Miura T, Masago Y, Konta Y, Tohma K, Manaka T, et al. Environmental surveillance of norovirus genogroups I and II for sensitive detection of epidemic variants. Appl Environ Microbiol. 2017;83(9).

18. Markt R, Stillebacher F, Nägele F, Kammerer A, Peer N, Payr M, et al. Expanding the Pathogen Panel in Wastewater Epidemiology to Influenza and Norovirus. Viruses. 2023;15(2):263.

19. Hellmér M, Paxéus N, Magnius L, Enache L, Arnholm B, Johansson A, et al. Detection of pathogenic viruses in sewage provided early warnings of hepatitis A virus and norovirus outbreaks. Appl Environ Microbiol. 2014;80(21).

20. Flood MT, D’Souza N, Rose JB, Aw TG. Methods Evaluation for Rapid Concentration and Quantification of SARS-CoV-2 in Raw Wastewater Using Droplet Digital and Quantitative RT-PCR. Food Environ Virol. 2021;13(3).

21. Haramoto E, Kitajima M, Kishida N, Konno Y, Katayama H, Asami M, et al. Occurrence of pepper mild mottle virus in drinking water sources in Japan. Appl Environ Microbiol. 2013;79(23).

22. Whale AS, De Spiegelaere W, Trypsteen W, Nour AA, Bae YK, Benes V, et al. The Digital MIQE Guidelines Update: Minimum Information for Publication of Quantitative Digital PCR Experiments for 2020. Clin Chem. 2020;66(8).

23. Tarkoma S, Alghnam S, Howell MD. Fighting pandemics with digital epidemiology. Vol. 26, EClinicalMedicine. 2020.

24. Arabzadeh R, Grünbacher DM, Insam H, Kreuzinger N, Markt R, Rauch W. Data filtering methods for SARS-CoV-2 wastewater surveillance. Water Science and Technology. 2021;84(6).

25. Havumaki J, Eisenberg JNS, Mattison CP, Lopman BA, Ortega-Sanchez IR, Hall AJ, et al. Immunologic and Epidemiologic Drivers of Norovirus Transmission in Daycare and School Outbreaks. Epidemiology. 2021;32(3).

26. Fitzgerald SF, Rossi G, Low AS, McAteer SP, O’Keefe B, Findlay D, et al. Site Specific Relationships between COVID-19 Cases and SARS-CoV-2 Viral Load in Wastewater Treatment Plant Influent. Environ Sci Technol. 2021;55(22).

