## Appendix for "Norovirus GII wastewater monitoring for epidemiological surveillance"

### Supplementary Figures

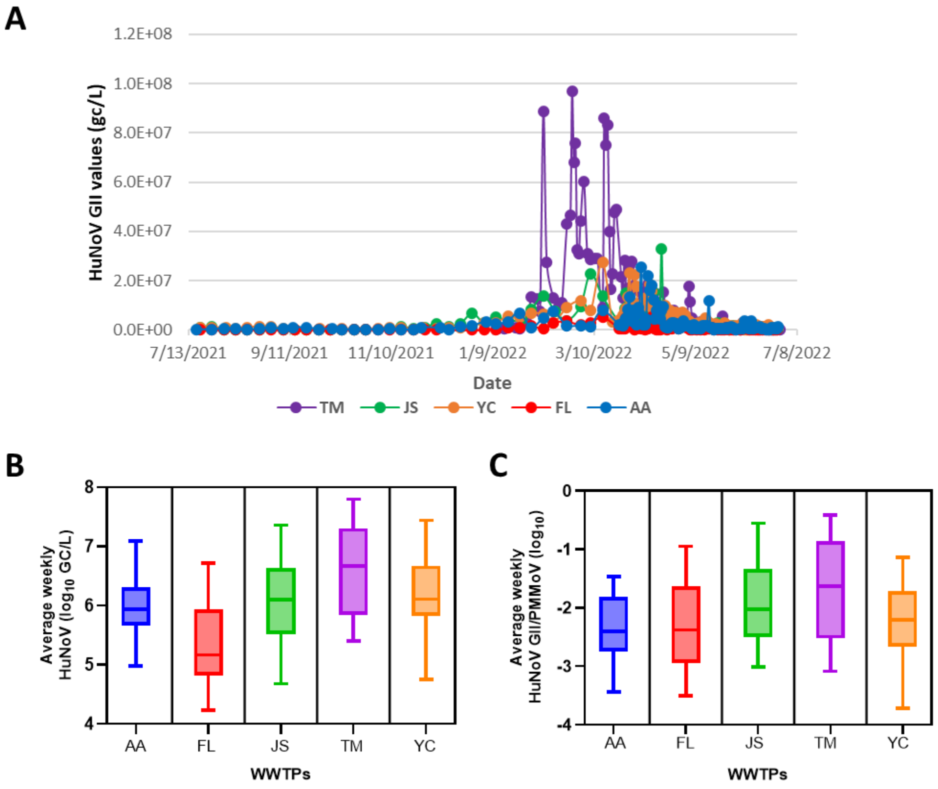


**Supplementary Figure 1. Comparison of HuNoV GII levels from 5 WWTPs in Michigan from 2021-2022.** HuNoV was quantified in influent samples using ddPCR at least weekly and gene copies per liter (gc/L) were plotted over one year. The only exception is TM, where sample collection began later, in January 2022. B and C. Box-and-whisker plots of log10 (B) average weekly gc/L HuNoV values (median, IQR) or (C) HuNoV/PMMoV values for each WWTP for the entire test period.

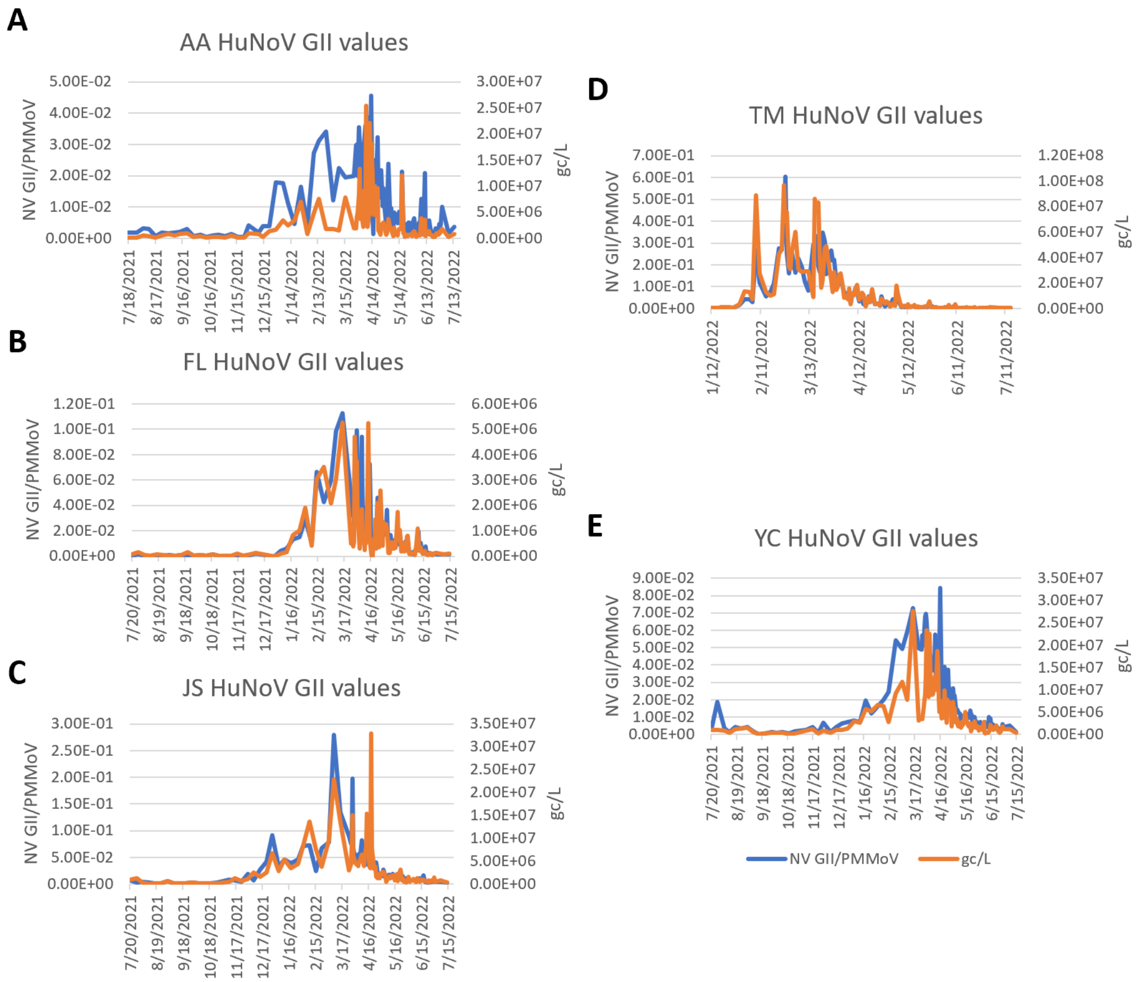


**Supplementary Figure 2. Individual graphs of HuNoV GII levels from 5 WWTPs in Michigan from 2021-2022 analyzed by two different methods.** HuNoV was quantified in influent samples using ddPCR at least weekly and gene copies per liter (gc/L), shown in orange, as well as HuNoV values normalized to the fecal indicator PMMoV, shown in blue, were plotted over time. The only exception is TM, where sample collection began later, in January 2022. Values from WWTPs in A. Ann Arbor (AA), B. Flint (FL), C. Jackson (JS), D. Tecumseh (TM), and E. area around Ypsilanti Community (YC).

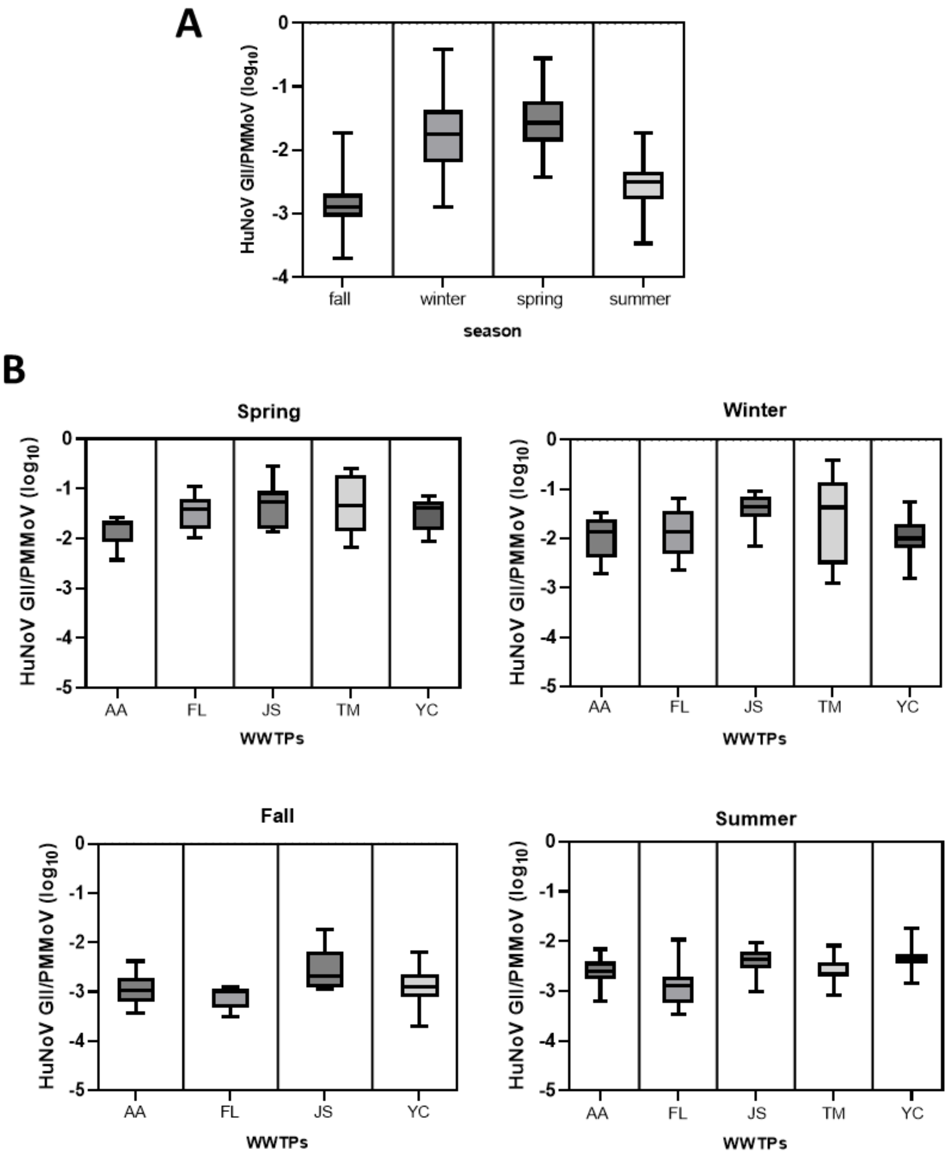


**Supplementary Figure 3.** Seasonal variations in PMMoV-normalized HuNoV GII levels in wastewater. A. HuNoV was quantified in influent samples using ddPCR at least weekly. The only exception is TM, where sample collection began later, in January 2022. The weekly averages of HuNoV/PMMoV were analyzed and a box and whisker plot were used to display the $log_{10}$ values (median, IQR). B. Seasonal HuNoV GII values (median, IQR) for each individual WWTP are presented using box-and-whisker plots. Note, no fall values for TM were obtained. The definition of the seasons is meteorological, beginning on the 1st day of the equinoxes or solstices.

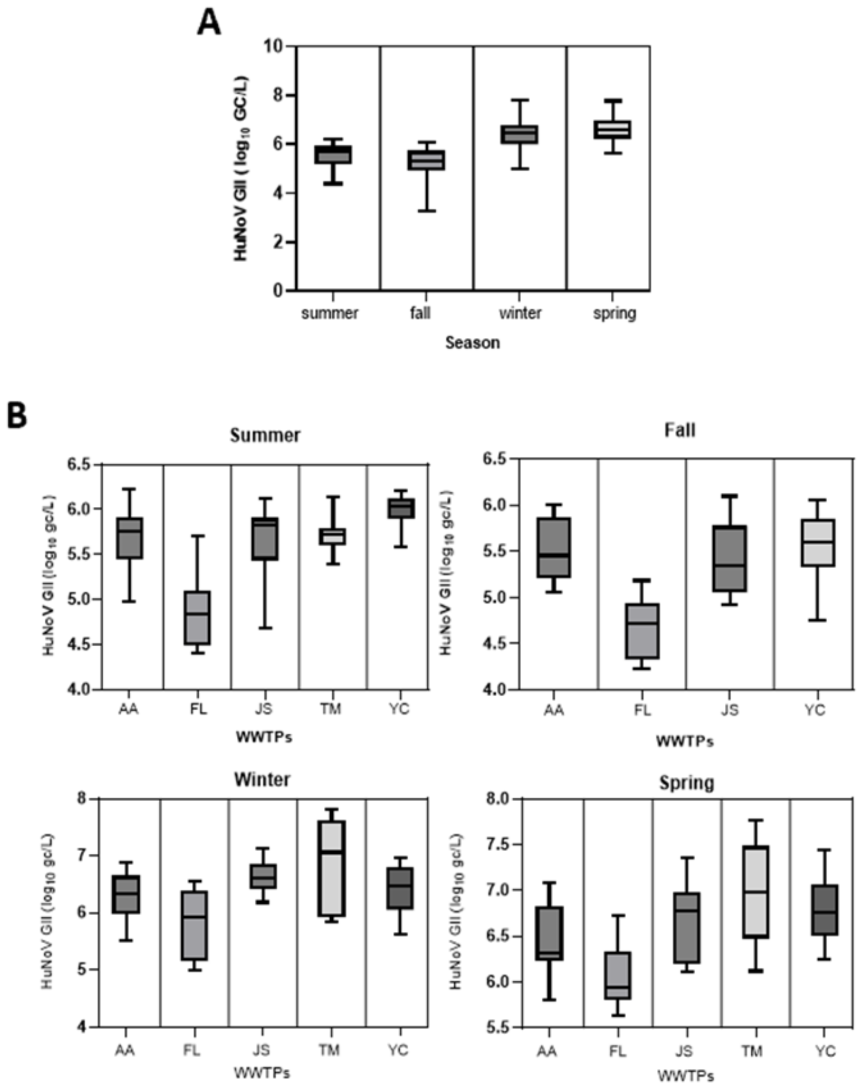


**Supplementary Figure 4.** Seasonal variations in HuNoV GII levels (gc/L) in wastewater. A. HuNoV was quantified in influent samples using ddPCR at least weekly. The only exception is TM, where sample collection began later, in January 2022. The weekly averages of HuNoV GII GC/L were analyzed and a box and whisker plot were used to display the $log_{10}$ values (median, ICR). B. Seasonal HuNoV GII values (median, IQR) for all WWTPs are presented using box-and-whisker plots. Note, no fall values for TM were obtained.

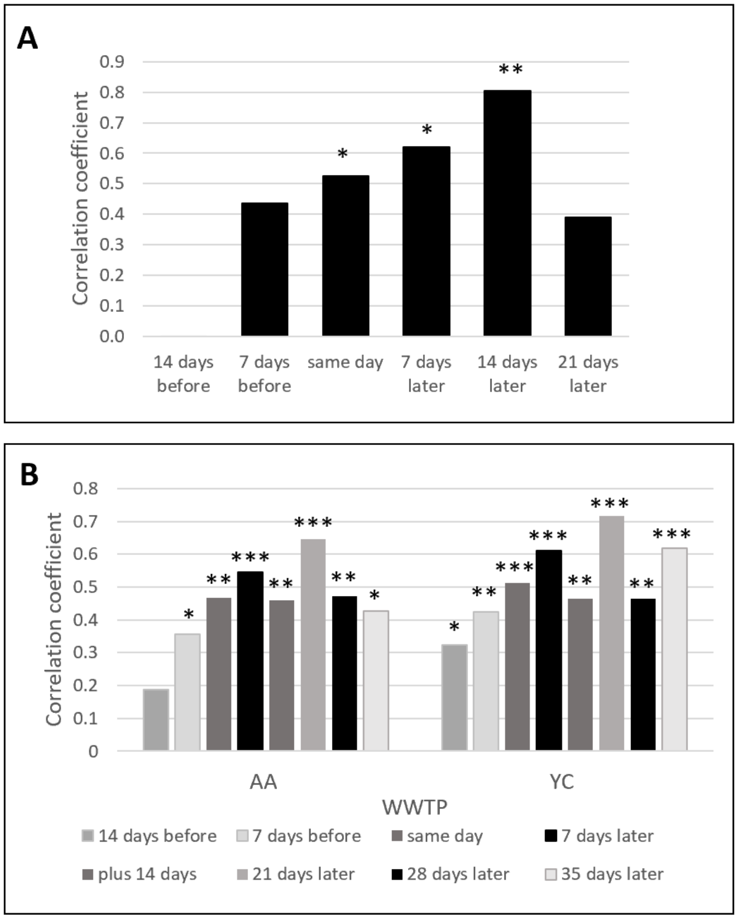


**Supplementary Figure 5.** Cross correlations of HuNoV GII wastewater values in gc/L with weekly school reported gastrointestinal illnesses. Graphs show Pearson's correlation coefficients (r) and probability values (p) determined by comparing weekly average wastewater data in HuNoV II gc/L, to school reported GI illnesses. A. Weekly number of school reported GI illnesses in TM schools was compared to weekly average TM WWTP HuNoV GII values. B. Weekly County-level school reported GI illness values normalized for attendance were compared to weekly average WWTP HuNoV GII values for AA and YC. Cross correlations were tested for a lead (14 or 7 days before), same timing, or a lag (7, 14, 21 and in some cases 28 and 35 days later) in school reported GI illnesses compared to HuNoV GII values. Note: * = $p<0.05$, ** = $p<0.01$, and *** = $p<.001$.

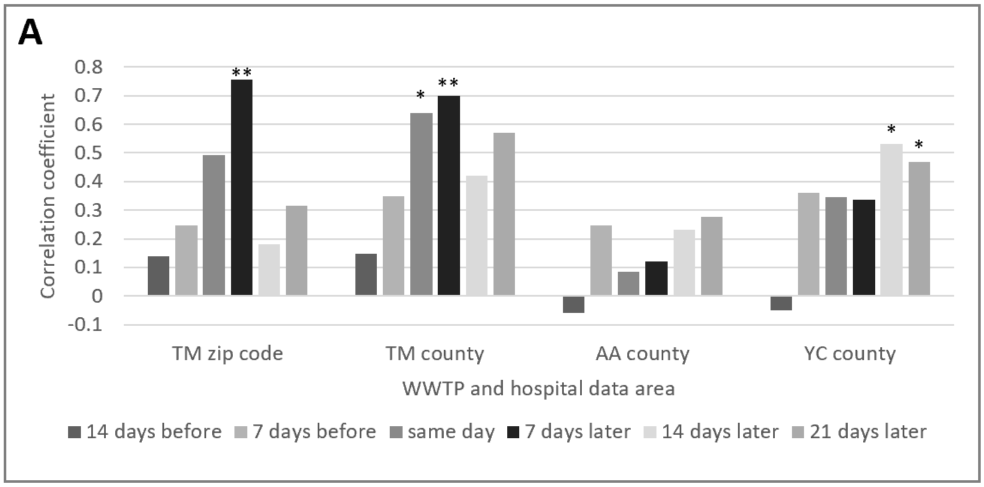


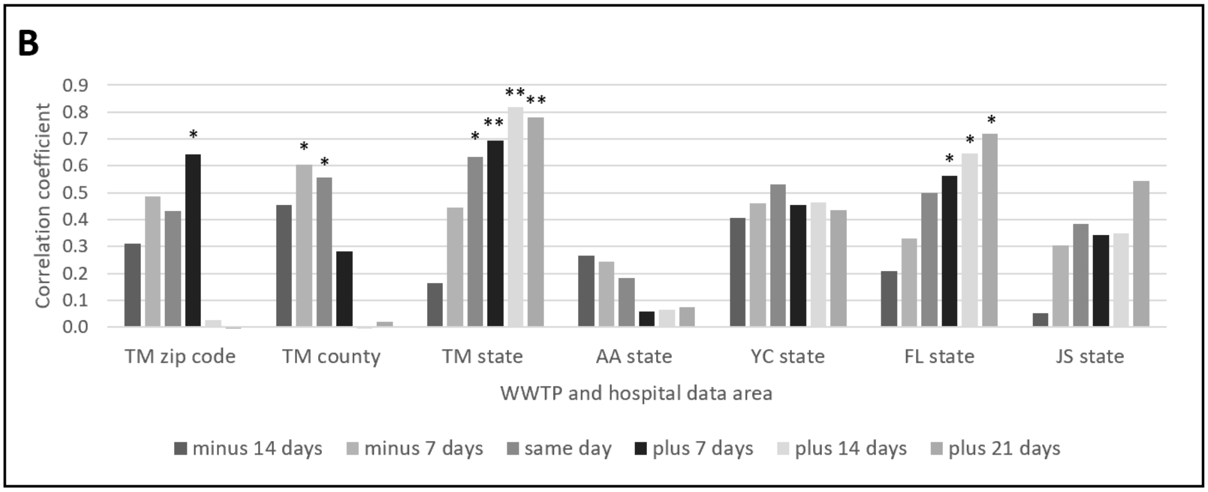


**Supplementary Figure 6. Correlation of HuNoV GII gc/L wastewater values with weekly GI related emergency department visits at regional and state levels. A**. Graph showing Pearson's correlation coefficients (r) and probability values (p) determined by comparing HuNoV GII wastewater values (in gc/L) from the TM, AA, and YC WWTPs to the weekly total GI related emergency department visits reported for patients from the corresponding counties (Lenawee and Washtenaw), and the TM zip code, for January – April 2022. **B.** Graph showing Pearson's correlation coefficients (r) and probability values (p) determined by comparing wastewater data as in A, to the weekly percentage of GI related emergency department visits compared to total visits. Data was reported for patients from the TM WWTP zip code and associated county (Lenawee). State (MI) hospital values were also compared to HuNoV GII wastewater values from all 5 WWTPs for January – April 2022. Cross correlations were tested for a lead (14 or 7 days before), same timing, or a lag (7, 14, and 21 days). Note: * = $p<0.05$, ** = $p<0.01$, *** = $p<0.001$

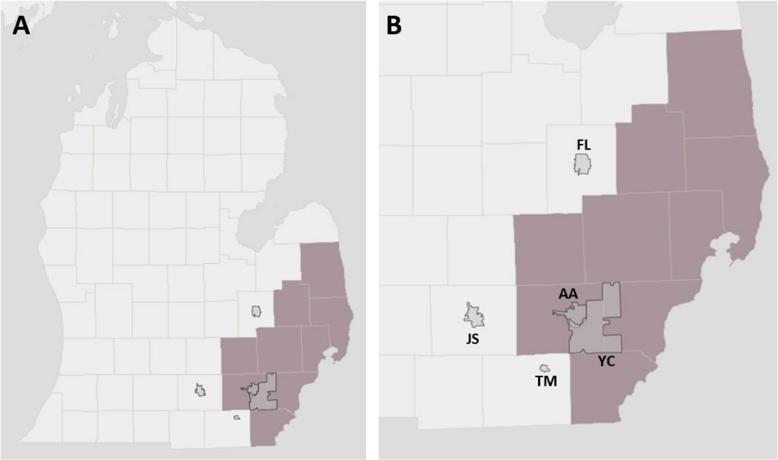


**Supplementary Figure 7. Area contained in google search trends dataset.** A. Map showing the lower peninsula of Michigan. B. A zoomed in version of southeast Michigan with the region included in the Detroit Metro google search trend dataset in mauve. Each of our WWTP catchment areas is shown in gray and labeled.

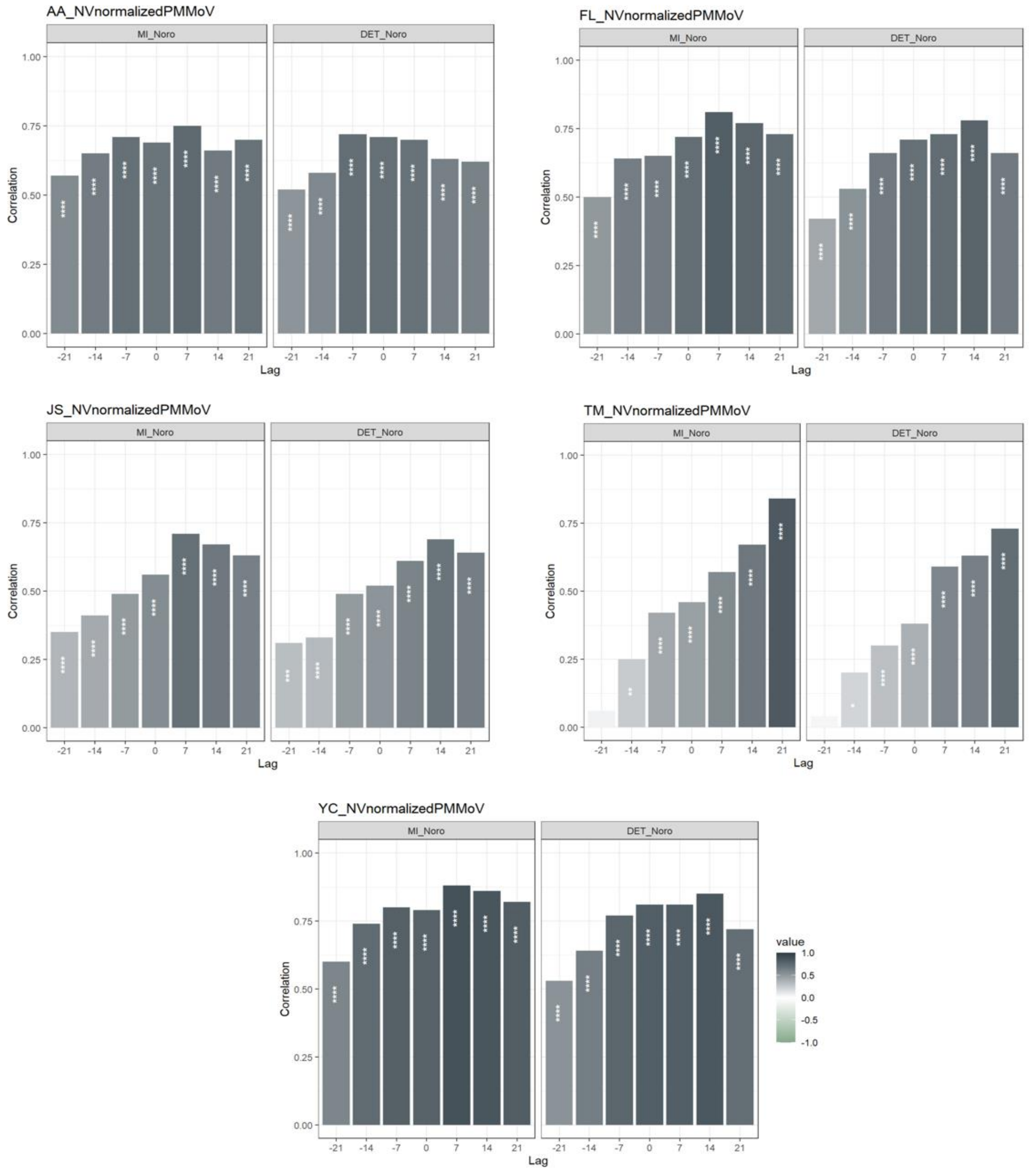


**Supplementary Figure 8.** Cross correlations comparing HuNoV GII/PMMoV values from 5 WWTPs and search term trends for "Norovirus" in the state of Michigan (left) and the Detroit Metro area (right). Pearson's correlation coefficients (r) and probability values (p) were determined. Cross correlations were tested for a lead (-21, -14, -7 days), same timing, or a lag (7, 14, 21, days) in search term trends compared to HuNoV GII/PMMoV values. Note: * = p<0.05, ** = p<0.01, *** = p<.001

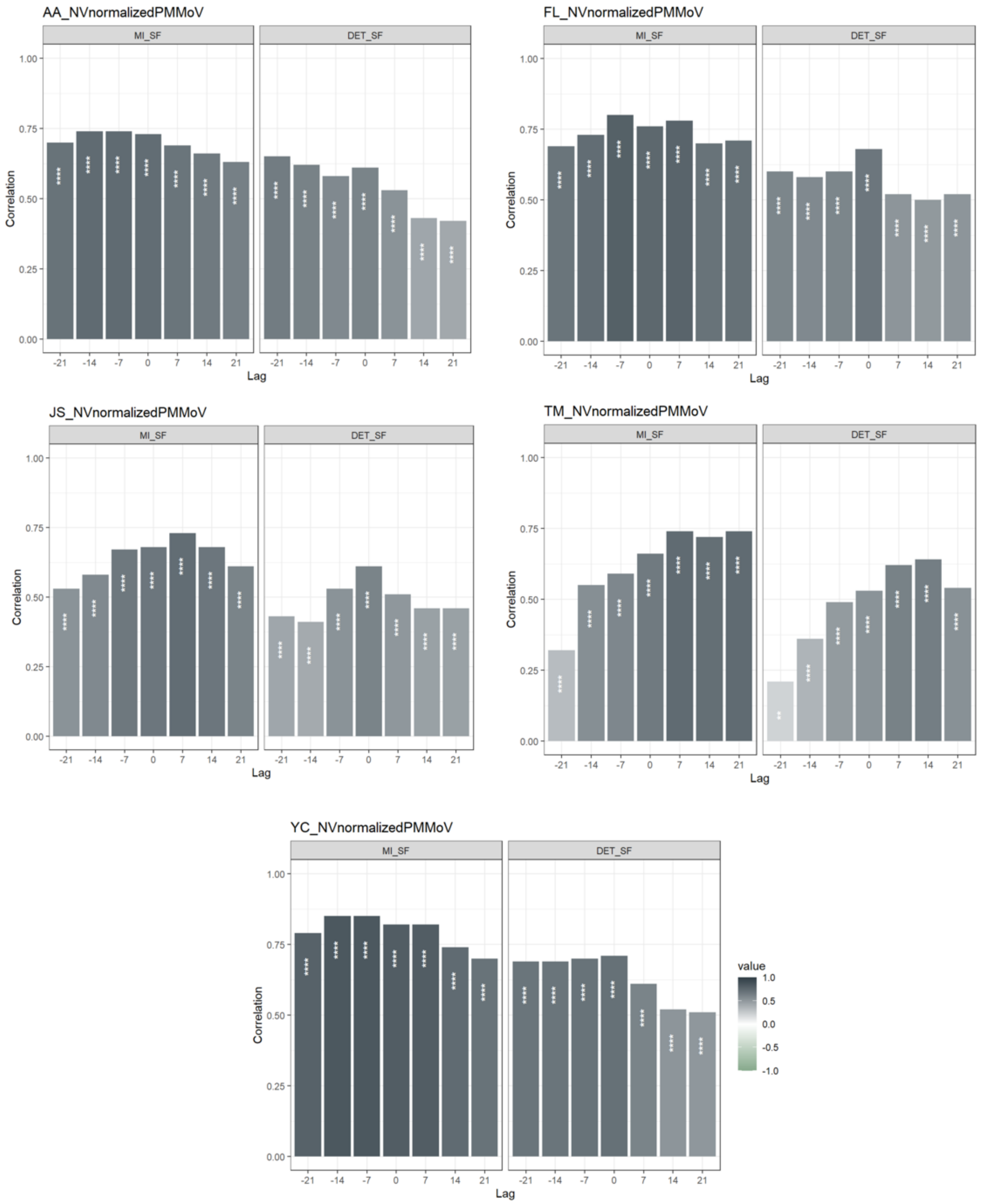


**Supplementary Figure 9.** Cross correlations comparing HuNoV GII/PMMoV values from 5 WWTPs and search term trends for "Stomach flu" in the state of Michigan (left) and the Detroit Metro area (right). Pearson's correlation coefficients (r) and probability values (p) were determined. Cross correlations were tested for a lead (-21, -14, -7 days), same timing, or a lag (7, 14, 21, days) in search term trends compared to HuNoV GII/PMMoV values. Note: * = p<0.05, ** = p<0.01, *** = p<.001

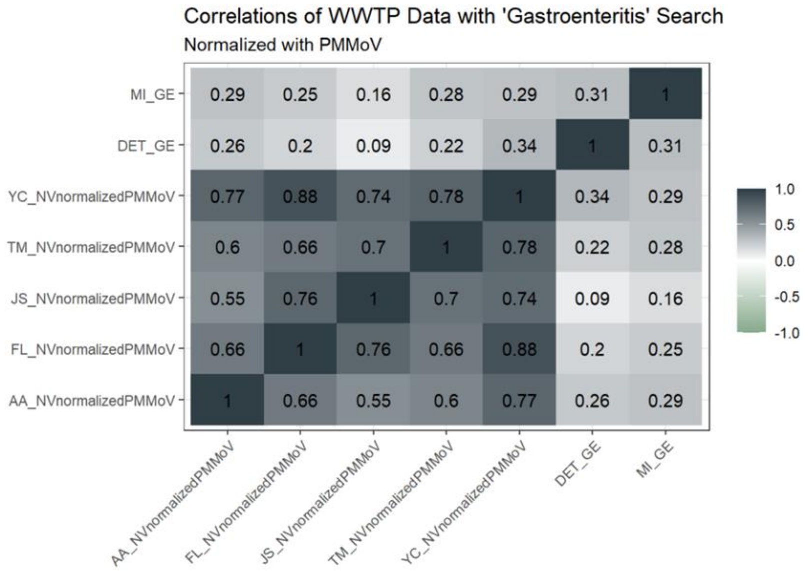


**Supplementary Figure 10.** Correlations were performed to determine the Pearson's correlation coefficients (r) for HuNoV GII/PMMoV values for all 5 WWTPs (AA, FL, JS, TM, and YC) with search term trends for "Gastroenteritis" (GE) in the Detroit Metro (Det Met) area and Michigan (MI).

**<u>Supplementary information</u>**

Including all 5 WWTPs, the daily values ranged from 6 x $10^3$ gc/L to 9.7 x $10^7$ gc/L (Supplementary Figure 1A); and the median weekly value was 6.0 $\log_{10}$ gc/L, with an interquartile range (IQR) of 5.52 to 6.48. Tecumseh (TM) samples had the highest levels with a mean concentration of 6.61 log10 and Flint (FL) samples had the lowest levels with a mean concentration of 5.36 log10 (Supplementary Figure 1B). The PMMoV normalized median weekly value for all 5 plants was -2.19 log10 (IQR = -2.73 to -1.59), and all 5 individual WWTPs including FL, have median values that fall in the interquartile range for the entire group (Supplementary Figure 1C).

**Supplementary methods**

### RT-ddPCR

The ability to monitor SARS-CoV2 levels in wastewater has resulted in a large increase in wastewater-based epidemiology research and has provided a means to establish methods of obtaining quantitative data for other pathogens that can be used to inform public health decisions. Norovirus GII had previously been shown to be detectable in WWTP influent (1,2).

Reverse transcription - digital droplet PCR (RT-ddPCR) analysis of Norovirus GII was performed on new RNA samples using multiplexing (with SARS N1 and N2) or triplexing (with BCoV and PMMoV) from March 25, 2022 to July 14, 2022. RNA from samples July 18, 2021 to March 25, 2022 were freeze-thawed once prior to ddPCR, and had been stored at -80°C for 1-8 months. A total of 640 samples were tested. Gene copies were quantified through one-step ddRT-PCR (n=3) using the One-step RT-ddPCR Advanced kit for Probes (catalog #1864021, Bio-Rad, CA) with the method described by Flood et al with some modifications (3). RT-ddPCR reactions were run at 50C for 60 minutes, 95C for 10 minutes, 40 cycles of 95C for 30 seconds and 56C for 1 minute, 98C for 10 minutes, and held at 4C. HuNoV GII-specific primers and probes (synthesized by Integrated DNA Technologies, IA) that target a 97bp region of the ORF1-ORF2 region and were previously tested for ddPCR were used (4,5). When Norovirus GII levels were low, 5$\mu$l undiluted RNA was analyzed by multiplexing with the SARS-CoV-2 nucleocapsid 1 (N1) and nucleocapsid 2 (N2) gene targets designed by the US Centers for Disease Control and Prevention (CDC) (6) using the 5'-FAM tagged Norovirus GII probe at half the concentration of the N1 and N2 probes (125nM for the norovirus probe, 250nM for SARS-CoV2 probes). When Norovirus GII levels were high, samples were diluted 1:100 and 5$\mu$l of the diluted RNA was analyzed by triplexing with BCoV and PMMoV targets using both 5'-FAM and 5'-HEX Norovirus GII probe at half the concentration of the BCoV and PMMoV probes (125nM for both norovirus probes, 250nM for BCoV and PMMoV probes). Primer concentrations were all 900nM. All sample and control reactions were run in triplicate. Droplet analysis was performed on the Bio-Rad QX200 droplet digital PCR systems (Bio-Rad, CA) with explanations of thresholding provided in Supplementary Figures 11 and 12. The results from replicate wells were merged. Controls were run along with all PCR reactions and included non-template controls, extraction controls, and positive controls. The N1/N2 primer/probe stocks and positive PCR controls were provided by MSU Rose lab as part of the SEWER project. For the Norovirus GII PCR control, two different controls were used. A 390 bp G-block (IDT) DNA, specific to the ORF1-2 region, was used as a PCR control. Additionally, non-infectious intact Norovirus GI and GII particles (Cat# NATNOV-6MC, Zeptometrix, Buffalo, NY) were used to confirm the efficiency of norovirus RNA extraction and the specificity of the HuNoV GII primer/probe set in ddPCR reactions. A 50-fold dilution series of HuNoV GII was added to both water and influent prior to performing ddPCR to confirm the gene copy range and lack of inhibition in the reactions. Plates containing negative and no template control wells that had greater than 3 droplets were rerun or re-extracted, consistent with the protocol used by the SEWER network for SARS-CoV2 ddPCR. Samples with less than 30% of the control reaction BCoV values were flagged for further analysis to determine if RNA degradation had occurred and re-extracted from PEG concentrates when appropriate. All primers, probes, and the Norovirus GII DNA control used are listed in Supplementary Table 1 (4,6–9). The limit of detection was established by serial dilution of the Norovirus GII DNA control and taking into account the limit of the blank (up to 3 positive droplets). Due to differences in sample concentration volumes after PEG precipitation (from 1-5mls) the limit of detection varies from $6 \times 10^3$ to $2.4 \times 10^4$ gc/L.

**Supplementary Table 1.** Primers, probes, and positive control used in RT-ddPCR.

| Target | Primer/Probe | Sequence | Reference |
|---|---|---|---|
| SARS-CoV-2 N1 | 2019-nCoV_N1-Fwd | 5’-GACCCCAAAATCAGCGAAAT-3’ | 6 |
| | 2019-nCoV_N1-Rev | 5’-TCTGGTTACTGCCAGTTGAATCTG-3’ | |
| | 2019-nCoV_N1-P | 5’-FAM-ACCCCGCATTACGTTTGGTGGACC-BHQ1-3’ | |
| SARS-CoV-2 N2 | 2019-nCoV_N2-Fwd | 5’-TTACAAACATTGGCCGCAAA-3’ | 6 |
| | 2019-nCoV_N2-Rev | 5’-GCGCGACATTCCGAAGAA-3’ | |
| | 2019-nCoV_N2-P | 5’-HEX-ACAATTTGCCCCCAGCGCTTCAG-BHQ1-3’ | |
| BCoV | BCoV-Fwd | 5’-CTGGAAGTTGGTGGAGTT-3’ | 7 |
| | BCoV-Rev | 5’-ATTATCGGCCTAACATACATC-3’ | |
| | BCoV-P | 5’-CCTTCATATCTATACACATCAAGTTGTT-3’ (5' FAM/ZEN/3' IBFQ) | |
| PMMoV | PMMoV-Fwd | 5’-GAGTGGTTTGACCTTAACGTTTGA-3’ | 8,9 |
| | PMMoV-Rev | 5’-TTGTCGGTTGCAATGCAAGT-3’ | |
| | PMMoV-P | 5’-CCTACCGAAGCAAATG-3’(5' HEX/ZEN/3' IBFQ) | |
| HuNoV GII | NV GII -Fwd | 5’-CAGGAGCCTATGTTCAGATGGATGAG-3’ | 4 |
| | NV GII -Rev | 5’-TCGACGCCATCTTCATTCACA-3’ | |
| | NV GII -P | 5’-TGGGAGGGCGATCGCAATCT-3’ (5' HEX or FAM/ZEN/3' IBFQ) | |
| HuNoV GII | | GACAAGAGCC AATGTTCAGA TGGATGAGAT TCTCAGATCT GAGCACGTGG GAGGGCGATC GCAATCTGGC TCCCAGTTTT GTGAATGAAG ATGGCGTCGA GTGACGCCAA CCCATCTGAT GGGTCCGCAG CCAACCTCGT CCCAGAGGTC AACAATGAGG TTATGGCTCT GGAGCCCGTT GTTGGTGCCG CCATTGCGGC ACCTGTAGCG GGCCAACAAA ATGTAATTGA CCCCTGGATT AGAAACAATT TTGTACAAGC CCCTGGTGGA GAGTTTACAG TATCCCCTAG AAACGCTCCA GGTGAAATAC TATGGAGCGC GCCCTTGGGA CCTGATCTAA ATCCCTACCT ATCCCATTTG GCCAGAATGT ACAATGGTTA TGCAGGTGGT | This paper |

### Examples of RT-ddPCR analysis

The ddPCR reactions were analyzed using the QuantaSoft Analysis Pro software (Bio-Rad, CA) in both one dimension and two dimensions. Examples are provided in Supplementary Figures 11 and 12 below.

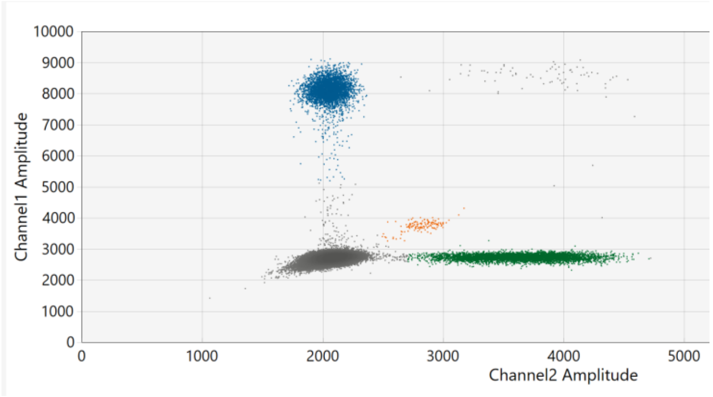


**Supplementary Figure 11.** Two-dimensional analysis of triplex reactions with BCoV, HuNoV GII, and PMMoV using samples diluted 1:100. The BCoV cloud in channel 1 was detected around 7500-9000 (blue), PMMoV cloud in channel 2 was detected from around 2800 to 4500 (green), and HuNoV GII cloud was detected between 3500 and 4000 in channel 1 and 2500 to 3000 in channel 2.

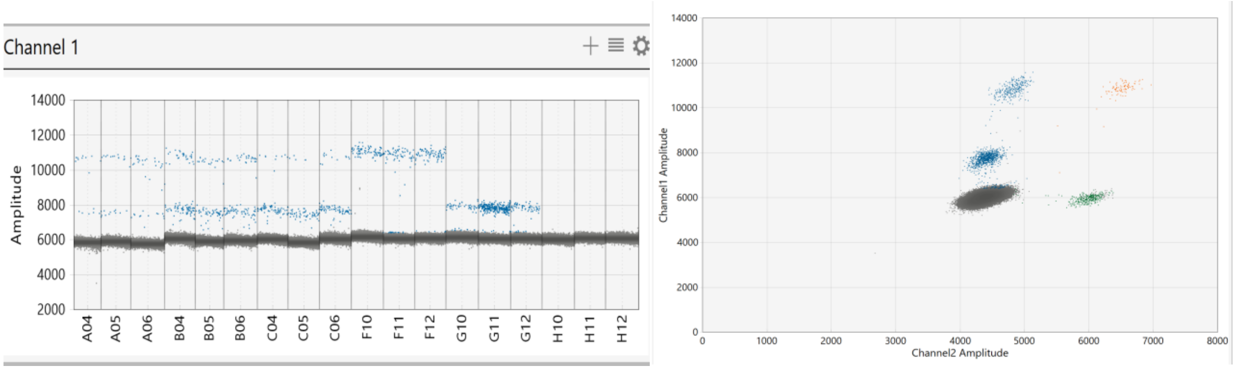


**Supplementary Figure 12.** One-dimensional (left) and two-dimensional (right) analysis of N1 and HuNoV GII using undiluted samples.  HuNoV GII positive controls were run in lanes G10-G12, SARS N1 positive controls were run in F10-F12, and negative extraction controls were in H10-H12. SARS N1 was detected from 9,000-11,500 and HuNoV GII was detected from 7,000-8,500 in channel 1.

### Correlation and Cross Correlation Analyses with Public Health Surveillance Data

***Hospital data.*** The Washtenaw County Public Health Department provided an EpiAlert Report containing daily gastrointestinal (GI) related emergency department (ED) visits made by Washtenaw County residents (which includes AA and some of the YC WWTP population) from 1/1/2022 to 6/15/2022. For a case to be included in the gastrointestinal category, chief complaints from the emergency department (ED) visits would include: abdomen pain or cramping, nausea, vomiting, diarrhea, and abdominal distention or swelling. A temporal cross-correlation was performed using GI related ED visits -2 to +3 weeks after the average weekly AA and YC WWTP HuNoV GII values (in both gc/L and HuNoV GII/PMMoV). We determined the correlation coefficients of weekly total ED GI visits of residents of Washtenaw County to weekly average wastewater HuNoV values from the AA and YC WWTPs using Graphpad Prism 9.4.1. Cross correlations were performed in weekly intervals from -2 to +3 weeks, where +3 weeks would refer to a lag in the hospital case values compared to the WWTP HuNoV GII values. The Pearson correlation coefficient (r) and the two tailed p-value (p) were determined for each comparison set.

The Lenawee County Public Health Department also provided gastrointestinal (GI) related emergency department visit data for Michigan residents, Lenawee County residents, and residents of the Tecumseh (TM) zip code from 1/22/2022 to 4/19/2022. For all three classes of residents the daily percentage of visits that were GI related was provided. For residents from Lenawee County and the TM zip code, but not the state, the daily total number of ED visits that were GI related was also provided. Using the method described above, cross correlations were performed using weekly total ED GI related visits of residents of Lenawee County and residents of the TM zip code, to weekly average wastewater HuNoV values from the TM WWTP. Additionally, the weekly average percentage of visits that were GI related (state, county, and zip code) were compared to the average wastewater HuNoV values from the TM WWTP, as this was the only hospital data provided at the state level. The weekly average percentage of visits that were GI related in the state of Michigan was also compared to the average wastewater HuNoV values from all of the WWTPs from -2 to +3 weeks as described above for data from Washtenaw County.

***School data.*** The Washtenaw County Public Health Department provided Washtenaw County daily school attendance numbers and school-reported gastrointestinal (GI) illnesses from September 26, 2021 through July 9, 2022. Due to the large variations in Washtenaw County school attendance, we normalized GI reports for Washtenaw County per 1000 students in attendance. We note this normalization is somewhat complicated as students with GI illness may not attend school, however it allows us to normalize for the broad changes in school population seen over time due to breaks/holidays/etc. Cross correlations were performed with Graphpad Prism 9.4.1 to determine Pearson's coefficients by comparing the Washtenaw County schools GI reports per 1000 students to weekly average wastewater HuNoV values from the AA and YC WWTPs from –2 to +4 weeks.

The Lenawee County Public Health Department also provided weekly total school-reported gastrointestinal (GI) illnesses for the TM schools from 1/1/2022 to 5/7/2022. Cross correlations were performed comparing the TM schools total weekly GI reports to weekly average wastewater HuNoV values from the TM WWTP from -2 to +3 weeks. The number of students in the district in 2021-2022, which was 2,545, was obtained from the National Center for Education Statistics website, although daily attendance values were not available.

### References

1. Kitajima M, Cruz MC, Williams RBH, Wuertz S, Whittle AJ. Microbial abundance and community composition in biofilms on in-pipe sensors in a drinking water distribution system. Science of the Total Environment. 2021;766.

2. Eftim SE, Hong T, Soller J, Boehm A, Warren I, Ichida A, et al. Occurrence of norovirus in raw sewage–a systematic literature review and meta-analysis. Water Res. 2017;111:366–74.

3. Flood MT, D'Souza N, Rose JB, Aw TG. Methods Evaluation for Rapid Concentration and Quantification of SARS-CoV-2 in Raw Wastewater Using Droplet Digital and Quantitative RT-PCR. Food Environ Virol. 2021;13(3).

4. Kageyama T, Kojima S, Shinohara M, Uchida K, Fukushi S, Hoshino FB, et al. Broadly reactive and highly sensitive assay for Norwalk-like viruses based on real-time quantitative reverse transcription-PCR. J Clin Microbiol. 2003;41(4).

5. Persson S, Eriksson R, Lowther J, Ellström P, Simonsson M. Comparison between RT droplet digital PCR and RT real-time PCR for quantification of noroviruses in oysters. Int J Food Microbiol. 2018;284.

6. Lu X, Wang L, Sakthivel SK, Whitaker B, Murray J, Kamili S, et al. US CDC real-time reverse transcription PCR panel for detection of severe acute respiratory syndrome Coronavirus 2. Emerg Infect Dis. 2020;26(8).

7. Decaro N, Elia G, Campolo M, Desario C, Mari V, Radogna A, et al. Detection of bovine coronavirus using a TaqMan-based real-time RT-PCR assay. J Virol Methods. 2008;151(2).

8. Haramoto E, Kitajima M, Kishida N, Konno Y, Katayama H, Asami M, et al. Occurrence of pepper mild mottle virus in drinking water sources in Japan. Appl Environ Microbiol. 2013;79(23).

9. Zhang T, Breitbart M, Lee WH, Run JQ, Wei CL, Soh SWL, et al. RNA viral community in human feces: Prevalence of plant pathogenic viruses. PLoS Biol. 2006;4(1).
